# Antibiotic Prescribing Patterns at COVID-19 Dedicated Wards in Bangladesh: A Single Center Point-Prevalence Survey

**DOI:** 10.1101/2021.01.15.21249868

**Authors:** Md. Maruf Ahmed Molla, Mahmuda Yeasmin, Md. Khairul Islam, Md. Mohiuddin Sharif, Mohammad Robed Amin, Tasnim Nafisa, Asish Kumar Ghosh, Monira Parveen, Md. Masum Hossain Arif, Junaid Abdullah Jamiul Alam, Syed Jafar Raza Rizvi, KM Saif-Ur-Rahman, Arifa Akram, AKM Shamsuzzaman

## Abstract

There is a clear deficiency in antimicrobial usage data and ongoing stewardship programs both in government and private health care facilities in Bangladesh. As evidences are mounting regarding irrational and often unnecessary use of antibiotics during COVID-19 pandemic, a point prevalence survey (PPS) was conducted across COVID-19 dedicated wards in Dhaka Medical College and Hospital (DMCH). Antibiotic usage data were collected from 193 patients at different COVID-19 dedicated wards at DMCH between 21 May, 2020 and 10 June, 2020. Comparisons in antibiotic usage were made between different groups using Pearson chi-square and Fisher exact test. Factors associated with multiple antibiotic prescription were evaluated using binary logistic regression model.

On survey date all (100%) patients were receiving at least one antibiotic with 133 patients (68.91%) receiving multiple antibiotics. Overall, patients presenting with severe disease received more antibiotics on average. Third generation cephalosporin ceftriaxone (53.8%), meropenem (40.9%), moxifloxacin (29.5%) and doxycycline (25.4%) were the four most prescribed antibiotics among survey patients. Among comorbidities diabetes mellitus (DM) was independently associated with increased antibiotic prescribing. Abnormal C-reactive protein (CRP) and serum d-dimer were linked with higher odds of antibiotic prescribing among survey patients. Overall, prevalence of antibiotic prescribing in COVID-19 patients at DMCH was very high. This could be attributed to a lack of clear treatment protocol against COVID-19 till date as well as lack of modern laboratory facilities to support judicial antibiotic prescribing in Bangladesh. A well-functioning antibiotic stewardship program in Bangladesh is required to prevent an impending health crisis.

## Introduction

The COVID-19 pandemic has led to an unprecedented crisis on every aspect of healthcare system across the world. Most of the infected people presenting with mild to moderate symptoms like cough, fever and lung infiltrate resemble bacterial pneumonia whereas only 20% of affected people get severe infection and only 6% people who become critically ill require ICU support [1]. Despite of the viral origin of COVID-19 and lack of evidence of bacterial super infection in huge number of cases, physicians are often compelled to prescribe a plethora of antimicrobials due to lack of specific antiviral treatment and vaccine against SARS-CoV-2, difficulties in differentiating between bacterial pneumonia and COVID-19, and uncertainty regarding secondary bacterial infection [2,3]. Different studies revealed that 70% of hospitalized patients receive one or more antibiotics, whereas it is scaled up to 100% in ICU setting [2,3]. Excessive prescribing and overuse of antibiotics is notable during this pandemic that, in the long run, might complicate the existing battle against antimicrobial resistance (AMR) [4,5]. Thus far, data on hospital antibiotic consumption and prescribing pattern during the COVID-19 pandemic are sparse, especially in countries without a well-functioning antimicrobial stewardship program. Point prevalence survey (PPS) on antibiotic usages among hospitalized patients reflects the actual scenario of antibiotic prescribing, which will aid in strategic planning of antibiotic stewardship program in countries like Bangladesh, where there is widespread ignorance among general population, as well as prescribers including pharmacists and traditional healers, regarding antibiotic resistance and how it might impact their future disease course. This single center PPS conducted at DMCH will fill the knowledge gap and aid in fighting against antibiotic resistance in low and low-middle income countries.

## Materials and Methods

### Study design and participant selection

This single center point prevalence survey (PPS) was conducted at COVID-19 dedicated wards at Dhaka Medical College and Hospital (DMCH), one of the largest government run hospitals in Bangladesh, from May 31 to June 10. The study was carried out in accordance with WHO methodology for point prevalence survey on antibiotic use in hospitals.

Adult patients (≥18 years) with a confirmed SARS-CoV-2 PCR positive result were considered for this study. Suspected COVID-19 patients or patients awaiting their rt-PCR results were excluded from the study.

### Sample size calculation

Dhaka Medical College and Hospital, a 2300 bed tertiary level teaching hospital, is considered to be the largest medical institution in Bangladesh. Among those 883 beds are reserved for COVID-19 patients and, as of December 22, current occupancy at COVID-19 wards stand at 70%, with an average between 20-50% since the inauguration of COVID-19 dedicated wards. According to WHO guideline, hospitals with more than 800 inpatient beds, one out of three patients can be sampled and included within the survey. For this study, data were collected from a total of 227 COVID-19 patients. After excluding entries with missing data, 193 patients were included in final analysis.

### Clinical definition of COVID-19 patients

On admission patients were categorized in either mild/moderate/severe or critical group based on criteria set by WHO. For this study, patients at COVID-19 dedicated wards at DMCH were identified as suffering from either moderate or severe disease. The criteria set forward for clinical classification are as follows: for moderate disease patients present at emergency room with clinical signs of pneumonia (fever, cough, dyspnoea, fast breathing) but no signs of severe pneumonia, including SpO2 ≥ 90% on room air. As for severe disease, patients with SARS-CoV-2 positive PCR result present at emergency room with clinical signs of pneumonia (fever, cough, dyspnoea, fast breathing) plus one of the following: respiratory rate > 30 breaths/min; severe respiratory distress; or SpO2 < 90% on room air.

### Data regarding antimicrobials and other drug prescription

During initial hospital admission, patients were asked about history of antimicrobial consumption after onset of COVID-19 specific symptoms and before hospital admission, including much prescribed, over the counter drug Ivermectin and hydroxychloroquine. Since this study specifically deals with antibiotics, antimicrobials including anti-virals, anti-parasitic and anti-fungals were excluded from the final analysis. But topical applications, in accordance with WHO PPS guideline, including eye and ear drops were excluded from the antibiotic list. Patients were divided into two groups – patients with no or single antibiotic and patients with multiple antibiotics on their treatment sheets on the survey date.

Antibiotic dosage, indication for prescription, presence of bacterial co-infection and other microbiological investigations were not recorded, largely due to unavailability of data and limited resources.

### Co-morbidities and biochemical markers

Apart from demographic information co-morbidities including hypertension (HTN), diabetes mellitus (DM), ischemic heart disease (IHD), asthma, chronic obstructive pulmonary disease (COPD), chronic kidney disease (CKD), and pre-existing malignancy history were recorded. Patients were categorized in to two groups – patients with no or single comorbidity and patients with two or more comorbidities. Statistical association was sought between number of comorbidities, disease severity and antibiotic prescribing pattern.

Biochemical marker values, specifically ones associated with inflammatory conditions, such as neutrophil and lymphocyte percentage, C - reactive protein (CRP), d-dimer, and serum ferritin level were recorded from investigation files and were expressed as mean ± SD. Biochemical markers were loosely divided into two categories based on findings – normal and abnormal. Following values were considered as normal for aforementioned biochemical markers – neutrophil (45-70%), lymphocyte (20-40%), CRP (<10 mg/l), d-dimer (≤0.5 considered as negative screening), and serum ferritin (for men 24-336 µgm/l; for female 11-307 µgm/l). Value outside of normal range were grouped as abnormal biochemical findings.

### Data management and analysis

After collection of relevant data, anonymous datasets were sent to researchers tasked with statistical analysis. For this study, statistical analysis was done using statistical Package for the Social Sciences (SPSS) software version 25. Continuous variables were expressed as mean ± SD and categorical variables were expressed in numbers (n) and percentage (%). Fisher’s exact and Pearson’s chi-square tests were used to test association between variables.

Finally, age and sex adjusted binary logistic regression was performed to ascertain the likelihood of factors that may influence increased prescribing of antibiotics (≥2 antibiotics) among study patients. For all analysis, statistical significance was set at p<0.05.

### Ethical consideration

For this study, researchers collected all the data from patient history sheets and anonymous data were sent to core team for statistical analysis. No patient was interviewed during the study and hence informed written consent was waived. This study was approved by the ethical review board at Dhaka Medical College and Hospital.

## Results

### Demographic characteristics and disease severity categorization

For this survey study, 193 patients were enrolled, of whom 134 (69.4%) were male and 59 (30.6%) were female. Patient age ranged from 18 to 95 with mean age of participants being 50.44±14.08 years. Patients were either categorized as moderate or severe upon admission, with overall p-value being 0.128 (Table-1).

**Table 1:** Age distribution and clinical categorization of patients.

| Age Range | Frequency (n=193) | Moderate (n=89) | Severe (n=104) | p-value |
| --- | --- | --- | --- | --- |
| 18-25 | 11 (5.70%) | 7 (7.9%) | 4 (3.9%) | 0.146 |
| 26-35 | 22 (11.40%) | 12 (13.5%) | 10 (9.6%) |  |
| 36-45 | 39 (20.21%) | 16 (18%) | 23 (22.1%) |  |
| 46-55 | 51 (26.42%) | 27 (30.3%) | 24 (23.1%) |  |
| 56-65 | 48 (24.87%) | 22 (24.7%) | 26 (25%) |  |
| 66+ | 22 (11.40%) | 5 (5.6%) | 17 (16.4%) |  |

### Antibiotic prescribing pattern

All study patients (100%) received one or more antibiotics. In total, 193 study participants received a total of 389 antibiotics, with each patient receiving 2.01 antibiotics on average, from the time of hospital admission to survey date. Among them 60 patients (31.08%) received single antibiotic agent whereas remaining 133 patients (68.91%) received two or more antibiotics on the survey date. Ceftriaxone, a beta-lactamase stable broad-spectrum antibiotic, was found to be the highest prescribed drug with 104 patients (53.88%) out of 193 total participants receiving/having received the drug according to their treatment record (Table-2). Next in line, Meropenem, another broad-spectrum injectable antibiotic, were prescribed in 79 (40.9%) surveyed patients (Table-2). Doxycycline, an oral antibiotic, was prescribed in 49 patients (25.4%) (Table-2). All prescribed antibiotics recorded during this survey were of broad spectrum in nature. The relationship between prescribed antibiotics and disease severity is illustrated in table-2

**Table 2:** Antibiotics prescription and disease severity.

| Antimicrobial | Moderate (n=89) | Severe (n=104) | Total (n=193) | p-value |
| --- | --- | --- | --- | --- |
| **Ceftriaxone** | **55 (61.8%)** | **49 (47.11%)** | **104 (53.88%)** | **0.028** |
| **Meropenem** | **23 (25.8%)** | **56 (53.9%)** | **79 (40.9%)** | **<0.001** |
| Levofloxacin | 13 (14.6%) | 14 (13.5%) | 27 (14%) | 0.819 |
| Moxifloxacin | 22 (24.7%) | 35 (33.7%) | 57 (29.5%) | 0.177 |
| **Doxycycline** | **16 (18%)** | **33 (31.7%)** | **49 (25.4%)** | **0.029** |
| Azithromycin | 22 (24.7%) | 23 (22.1%) | 45 (23.3%) | 0.672 |
| Amoxicillin | 6 (6.7%) | 14 (13.5%) | 20 (10.4%) | 0.128 |

### Comorbidity, inflammatory markers, and disease severity

Results revealed a positive association between diabetes mellitus and number of antibiotics received per patient. Patients admitted into COVID-19 wards with diabetes mellitus as comorbidity were more likely to receive two or more antibiotics than patients admitted with other comorbidities (p=0.007) (Table-3). Besides, patients with multiple comorbidities were more likely to present with severe disease (p=0.005) but no statistical significance could be found between multiple comorbidities and increased antibiotic prescription (p=0.056) (Table-4).

**Table 3:** comorbidity and antimicrobial prescribing.

| Comorbidity | Antibiotic (0-1) | Antibiotic ($\geq 2$) | p-value |
| --- | --- | --- | --- |
|  | Yes/No (n=60) | Yes/No (n=133) |  |
| Hypertension | 25/35 | 70/63 | 0.160 |
| **DM** | **16/44** | **63/70** | **0.007** |
| IHD | 12/48 | 38/95 | 0.208 |
| Asthma | 13/47 | 20/113 | 0.258 |
| COPD | 5/55 | 14/119 | 0.636 |
| CKD | 3/57 | 11/122 | 0.417 |
| Malignancy | 0/60 | 2/131 | 0.474* |

\*Analyzed using 1-tail Fisher's exact test

**Table 4:** Association between total number of comorbidities and disease severity, antibiotic prescription.

| | Total comorbidity<br>(0-1) | Total comorbidity<br>($\geq 2$) | p-value |
| --- | --- | --- | --- |
| Antibiotic (0-1) (n=60) | 36 | 24 | 0.056 |
| Antibiotic ($\geq 2$) (n=133) | 60 | 73 | |
| Moderate disease (n=89) | 54 | 35 | **0.005** |
| Severe disease (n=104) | 42 | 62 |  |

Among the common inflammatory markers such as neutrophil and lymphocyte percentage, serum ferritin level, CRP, and d-dimer level, only CRP (p=0.005) and d-dimer (p=0.002) showed positive association with antibiotic prescription among study patients (Table-5). Additionally, there was significant correlation between disease severity and number of antimicrobials received per patient (p=<0.00001).

**Table 5:** Biochemical markers and antibiotic prescribing pattern.

| Biochemical marker | Mean±SD value | Antimicrobial number (0-1)<br>Normal/Abnormal | Antimicrobial number ($\geq 2$)<br>Normal/Abnormal | p-value |
| --- | --- | --- | --- | --- |
| Neutrophil | (76.8 ± 11.9) % | 16/44 | 35/98 | 0.959 |
| Lymphocyte | (18.3 ± 10.3) % | 21/39 | 43/90 | 0.715 |
| **CRP** | **(23.8 ± 27.2) mg/l** | **28/32** | **35/98** | **0.005** |
| **d-dimer** | **(1.3 ± 1.5) μgm/ml** | **29/31** | **34/99** | **0.002** |
| Serum ferritin | (630.8 ± 664.8) μgm/l | 17/43 | 31/102 | 0.455 |

On age and sex adjusted binary logistic regression model, significant statistical association was found between abnormal CRP and d-dimer status and prescription of ≥2 antibiotics. Patients categorized as having abnormal CRP were 2.32 (95% CI: 1.21-4.85) times more likely to receive≥2 antibiotics (p=0.025 on Wald test) (Table-6). Additionally, patients with abnormal d-dimer value were 2.13 (95% CI: 1.03-4.43) times more likely to receive ≥2 antibiotics (p=0.42 on Wald test) (Table-6).

**Table 6:** Age and sex adjusted binary logistic regression considering number of antibiotics received as dependent variable.

| Variables | p-value | aOR | 95% CI |  |
| --- | --- | --- | --- | --- |
|  |  |  | Lower | Higher |
| Age (Years) | 0.075 | 1.028 | 0.997 | 1.059 |
| Sex | 0.549 | 1.269 | 0.582 | 2.769 |
| Hypertension | 0.814 | .895 | 0.357 | 2.247 |
| Diabetes mellitus | 0.126 | 2.074 | 0.814 | 5.284 |
| Ischemic heart disease | 0.667 | .791 | 0.271 | 2.305 |
| Asthma | 0.743 | .850 | 0.321 | 2.249 |
| Chronic obstructive pulmonary disease | 0.781 | .828 | 0.218 | 3.137 |
| Chronic kidney disease | 0.605 | 1.481 | 0.334 | 6.557 |
| Total Comorbidity | 0.924 | .940 | 0.264 | 3.347 |
| Patient Condition | 0.218 | 1.554 | 0.771 | 3.131 |
| **CRP** | **0.025** | **2.325** | **1.114** | **4.849** |
| Serum Ferritin | 0.831 | 1.091 | 0.491 | 2.427 |
| **d-dimer** | **0.043** | **2.127** | **1.023** | **4.423** |
| Neutrophil | 0.663 | 0.789 | 0.273 | 2.283 |
| Lymphocyte | 0.775 | 0.866 | 0.322 | 2.328 |

## Discussion

Before the pandemic even began towards the end of 2019, antimicrobial resistance (AMR) and resistance to gram negative bacteria were considered as matter of concern on a global scale. There were already reports of widespread antibiotic resistances in previous reports with researchers undertaking innovative steps to address the issue in different settings across the globe [6,7]. To make matters worse, COVID-19 pandemic and the unavailability of a specific treatment to date have forced researchers and clinicians to resort to symptomatic management with reports of unnecessary antimicrobial prescription [8]. In a recent published study, majority (72%) hospitalized patients received antimicrobial treatment even though only 8% of them demonstrated bacterial or fungal co-infection [2]. Likewise, in another study conducted among ICU patients in 88 countries, 70% received one or more antibiotic or antifungal drug despite only 54% of them having proven bacterial co-infection [9]. This was the findings from a 2017 study. Considering the uncertainty regarding COVID-19 management these numbers may escalate during the pandemic.

As evidenced from few studies conducted on antimicrobial usage during the pandemic, in most cases antimicrobials were prescribed as prophylaxis or symptomatic treatment purpose without justified cause or supportive laboratory investigation. With growing reports of antimicrobial misusage, health care authorities around the globe asked for strict regulations regarding antimicrobial usage in COVID-19 patients [10,11]. Current guideline from WHO indicates that no antibiotic or antifungal drug should be prescribed in mild or moderate cases unless there are pre-existing symptoms of bacterial or fungal co-infection [8]. Furthermore, regarding empirical antimicrobial prescription in severe cases, patients’ overall health condition, local epidemiology and the clinical judgment from treating physician should be integrated to allow for judicial antimicrobial usage [8].

In Bangladesh, during the early months of pandemic, there were reports of widespread antimicrobial consumption among COVID-19 positive and suspected patients-in most cases even without a prescription from a certified physician. In a previous study conducted among Bangladeshi COVID-19 patients isolating at home with mild or asymptomatic infection, 63% received one or more antimicrobial agent including investigational drugs such as ivermectin, remdesivir and favipiravir [12]. This finding and overall global trend in antimicrobial usage necessitated a broader research on hospitalized patients in Bangladesh.

To date, this is probably the first and only PPS study in Bangladesh on antimicrobial usage in COVID-19 dedicated wards. Overall, the antimicrobial usage was very high with all hospital admitted patients with moderate or severe disease received at least one antibiotic agent. This number is very high compared to other published PPS study conducted in different countries. For example, PPS conducted in different hospitals in Scotland and Singapore during April, 2020 revealed at least one antimicrobial, which included antivirals and antifungals, was prescribed in 38.3% and 6.2% hospitalized SARS-CoV-2 positive patients respectively [13,14]. There are several possible factors behind such significant difference in antimicrobial prescribing pattern between Bangladesh and Scotland/Singapore. There is a well-functioning Scottish Antimicrobial Prescribing Group (SAPG), established in 2008, tasked with overseeing the pattern of antimicrobial usage in Scottish hospitals and implement antimicrobial stewardship programs in different hospitals [15]. Likewise, antibiotic prescription in Singapore hospitals is highly regulated by antimicrobial stewardship units and often supplemented by relevant laboratory investigations [16]. In contrast, there is no such governing body in Bangladesh, both in government and private hospitals, and antimicrobial usage in hospitals across the country is largely empirical. Furthermore, with a modest healthcare budget, it is tough for government run hospitals to perform modern biochemical tests such as procalcitonin, serum ferritin, CRP, d-dimer or send samples for microbiological culture for each and every hospital admitted patient [17].

Procalcitonin, a useful biomarker, used to predict bacterial co-infection and disease severity in COVID-19 patients, is costly and neither the patients nor the laboratories in Bangladesh at present can perform the test on regular basis [18]. Hence, physicians often have to rely solely on their clinical experience and provide symptomatic management.

Among all listed comorbidities diabetes mellitus was independently associated with increased antimicrobial usage. DM is a pre-infectious condition and previous studies suggest people admitted at hospitals with DM end up receiving more antibiotics than patients without DM, largely due to presence of antibiotic resistance in diabetic population [19]. This might have compelled physicians to prescribe multiple antibiotics to make sure the patient condition does not worsen. In contrast, in the Scottish study COPD was identified to be positively associated and diabetes mellitus was negatively associated with antimicrobial usage. Regarding the Singapore study, patients with multiple comorbidities were more likely to receive antibiotics. In this study, no such association could be established between multiple comorbidities and increased antibiotic prescription.

Third generation cephalosporin and meropenem were the two most prescribed drugs in Bangladeshi study whereas co-amoxiclav, amoxicillin and doxycycline were mostly prescribed in Scottish and Singapore hospitals. In addition, prevalence of multiple antibiotics prescription during the survey period was higher compared to the Scottish and Singapore study. This can be explained by the fact that most patients admitted in DMCH have little financial means to purchase drugs from outside. As a result, they often rely on hospital provided drugs and during the peak pandemic period, ceftriaxone and meropenem were supplied by the hospital at free of cost. Another valid explanation might be, all admitted patients in DMCH were either suffering from moderate or severe COVID-19 disease, and injectable antibiotics are mostly prescribed in such cases to prevent the condition from deteriorating further [2]. Additionally, inflammatory markers such as CRP and d-dimer were found to be positively associated with increased antibiotic prescribing among survey patients. This finding corroborates previous research findings where it was found that increased CRP and d-dimer is positively associated with poor patient outcome, possibly compelling physicians to prescribe multiple antibiotics in the process [20].

There are several limitations to this PPS study. Due to archaic method of data collection, storage and preservation in Bangladeshi hospitals, extracting meaningful data from patient history sheets were often impossible. Furthermore, the researchers did not record the route of administration, whether the patient was on oxygen therapy, or presence of co-infection in surveyed patients. Besides, this was a single center study. For a comprehensive outlook on antibiotic usage in Bangladeshi hospitals several other centers should have been included. Unfortunately, this was beyond the scope of this survey study. But future research into antimicrobial stewardship programs in Bangladeshi hospitals should take these shortcomings into consideration and perform a comprehensive analysis of overall antimicrobial usage situation in Bangladesh.

## Data Availability

If interested the readers can contact directly with the corresponding author for access to data and resources to replicate the findings discussed in this paper.

## Acknowledgements

The authors would like to acknowledge the contributions of Bangladeshi healthcare workers working at COVID-19 dedicated wards in different hospitals across the country.

